# “Social distancing: barriers to its implementation and how they can be overcome – a rapid systematic review”

**DOI:** 10.1101/2020.09.16.20195966

**Authors:** Mahan Sadjadi, Katharina Selda Mörschel, Mark Petticrew

**Author notes:** **Author Contributions:** MS and MP conceptualised the review and designed the search strategy. MS carried out the searches and selected studies for inclusion. KSM double checked that all selected studies met the inclusion criteria. MS and KSM appraised the quality of studies. MS extracted and analysed data, and synthesised review findings. KSM double coded and analysed a third of the studies, and synthesised findings together with MS. Review findings were discussed with MP. MS wrote the first draft, and all authors contributed to the final version of the manuscript. **Compliance with Ethical Standards**. **Conflict of interest statement:** The authors have no relevant financial or non-financial interests to disclose. **Role of funding source:** The authors did not receive support from any organization for the submitted work. **Ethical Approval:** Formal ethical approval was not required for this review.

## Abstract

**Aim:** To systematically review qualitative literature on social distancing in order to identify and describe factors that enable or prevent its implementation.

**Methods:** A rapid systematic qualitative review was conducted for which a comprehensive systematic search was carried out across eleven databases. Included papers report on primary qualitative studies of the barriers and facilitators to the implementation of social distancing measures in potentially epidemic infectious diseases. An adapted meta-ethnographical approach was used for synthesis. Review findings were assessed for strength and reliability using GRADE-CERQual.

**Results:** 29 papers were included from the systematic search that yielded 5620 results and supplementary methods. The review identifies two broad categories of barriers to social distancing measures: individual- or community-level psychological or sociological phenomena, and perceived shortcomings in governmental action. Based on this, 25 themes are identified that can be addressed to improve the implementation of social distancing.

**Conclusion:** There are many barriers, on different levels, to the implementation of social distancing measures. Among other findings, the review identifies the need for good communication as well as the need for authorities to provide comprehensive support as two key opportunities to increase acceptability and adherence. High-quality research is needed during the COVID-19 pandemic to better describe mechanisms by which implementation of social distancing can be improved, and, more importantly, what is already known has to be put into practice.

## Introduction

On 31 December 2019, the WHO was informed of an outbreak of pneumonia of unknown aetiology in the city of Wuhan, China (1). This was the starting point of a pandemic affecting millions of people. In the following weeks and months, as SARS-CoV-2 started to spread to an increasing number of countries, social distancing was rapidly established as a central part of containment efforts (2).

Social distancing measures are not new. They have been employed and researched previously, specifically during epidemics of diseases like SARS, MERS or pandemic forms of influenza (2–4). The modelling and observational studies that have been conducted suggest the important effect such measures can have, and with a lack of therapeutic options, decisive preventive action is necessary to be able to save lives (5–8).

In spite of some research around social distancing measures in the context of non-pharmaceutical interventions (4), it remains unclear what factors enable or prevent their implementation, and what determines their feasibility and acceptability in the eyes of the public that is expected to carry them out. This is a critical question because many of these measures depend on the participation of the whole population. Having a stronger understanding of what factors prevent or promote the implementation of and adherence to social distancing measures is crucial for designing an effective and ethical pandemic response.

To be able to provide guidance for policymaking and future research, this systematic qualitative review sets out to synthesise the evidence relating to factors that affect the implementation of social distancing measures.

## Methods

A rapid systematic review of qualitative research on social distancing was conducted. A protocol was outlined internally before the start of the review process. In order to ensure reflexivity in the conduct of this review, the lead reviewers considered, at the outset and throughout the review process, how their views and opinions were likely shaped by their first-hand experiences of social distancing implementation in Germany and the UK.

### Inclusion Criteria

Studies were included in this review if they:

a. reported on qualitative studies with primary data generation
b. addressed infectious diseases with human-to-human transmission and epidemic potential (Influenza, MERS, SARS, Ebola), and
c. included information on feasibility, acceptability, barriers, facilitators and attitudes regarding the implementation of social distancing measures.

### Search Strategy

#### The primary, defining search for “Social Distancing”

Despite the central role social distancing plays in the pandemic response, neither researchers nor policymakers or the media use consistent definitions. In order to build a search strategy that is sensitive to all measures that fall within the broad concept of social distancing, a primary, defining search was performed in MEDLINE, EMBASE, PsycINFO, Global Health, CINAHL and Cochrane Library databases for the search term “Social Distancing”. Additionally, websites and documents of the WHO (9,10), CDC (11,12), ECDC, China CDC and Africa CDC (13) were searched for definitions of social distancing. Searches were carried out on 13 March 2020.

The identified concepts for measures were policy-level interventions like mandated closure of schools, child-care facilities, restaurants, and public venues, the cancellation of public events, bans on public transportation as well as isolation and quarantine on the one hand, and individual-level behavioural responses, like workplace non-attendance, contact number reduction, staying home, avoiding crowds, avoiding transportation and reducing travel on the other hand.

The final search for qualitative studies on the acceptability, feasibility and implementation of social distancing measures was based on these findings and definitions.

### The final search

Based on the results of this primary investigation, a second search was performed that included all aspects of social distancing that were found through the first search. The general strategy was to combine terms related to social distancing with terms on mass gatherings, and to then combine those with terms around epidemics. In the end, a qualitative filter (developed by UThealth, https://libguides.sph.uth.tmc.edu/search_filters) was applied to the results. The full search strategy can be found in appendix 1. This final search was carried out between 17 and 19 March 2020 in MEDLINE, EMBASE, PsycINFO, Global Health, CINAHL, SCI-EXPANDED, SSCI, A&HCI, CPI-S, CPI-SSH, and ESCI. The most recent version of each database was used, and no time restrictions were applied.

### Study Selection

All the records retrieved were imported into Zotero 5.0 (https://www.zotero.org/download/) from which duplicates were removed and titles and abstracts were screened against the inclusion criteria. The selection of studies was discussed among the authors, and consensus was reached.

### Data Extraction

Data were extracted regarding the following aspects: setting, sample size and composition, data collection methods, study aims as well as first order (participant quotes) and second order themes (analysis and interpretation by study authors). This was done using a standardised form which was also used to synthesise third order meta-synthesis themes, and to track quality assessment.

### Quality Assessment

The quality of all included studies was assessed using the Critical Appraisal Skills Programme (CASP) assessment tool for qualitative studies. (14) The authors conducted their critical appraisal independently and discussed their assessments to reach consensus.

### Analytic Strategy and Synthesis

The review uses meta-ethnographical approaches adapted from Britten and colleagues (15).

Each paper was studied in-depth and themes that relate to the research question were identified inductively from the data. Line-by-line coding was done for relevant segments of reports. Participant statements quoted in research reports were treated as first order themes, and the analysis and interpretation by researchers were treated as second order themes. The third order meta-synthetical themes were formed inductively based on these previously identified themes following initial in vivo and subsequent axial coding. Differences between reviewers’ assessments were discussed until consensus was reached. The third-order themes were treated as the review’s findings. Confidence in each finding was assessed using the GRADE-CERQual approach, which considers methodological limitations, relevance, coherence, and adequacy of data (16). The quality assessment previously performed using CASP contributed to the weighing of study findings by informing the appraisal of the GRADE-CERQual “methodological limitations” category. Moreover, where themes were corroborated by multiple studies of which at least one was high-quality (defined has having no significant concerns regarding study design, recruitment, data collection and analysis, i.e. rated with “Yes”), overall minor quality concerns were reduced, and a high confidence rating was attributed for that finding. MS analysed all included studies and KM double-coded a third of the included studies. The authors reached consensus regarding identified themes and review findings.

### Reporting

This review follows PRISMA (17) and ENTREQ (18) statement guidelines.

## Results

### Description of search results and included studies

The final search (see figure 1) yielded 5620 results. After deduplication, 4019 titles and abstracts were screened. 147 papers which could not be excluded based on title and abstract remained for full text screening of which 28 papers were included. One additional paper was identified by searching references of studies.

**Figure 1:**
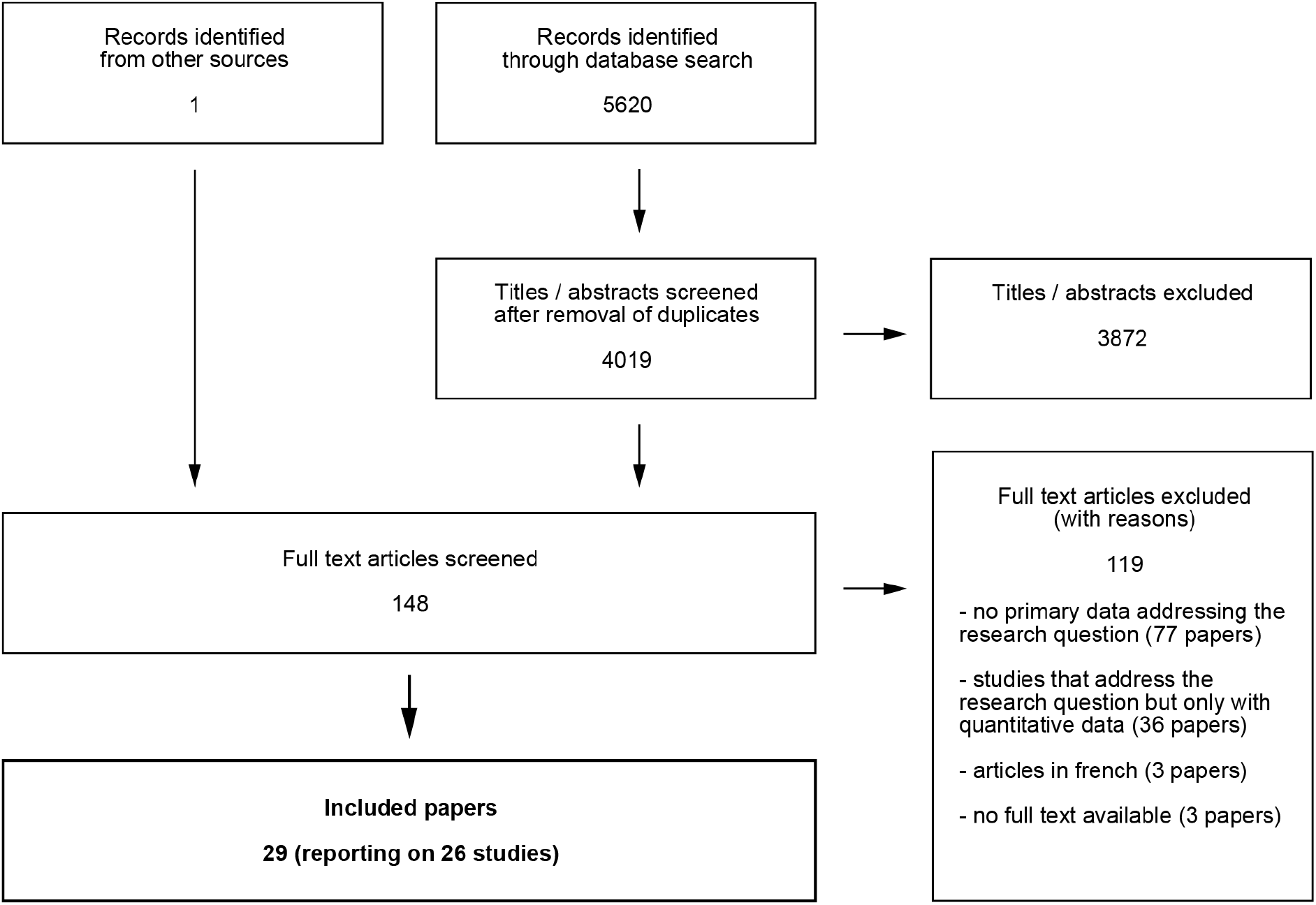
Flowchart for the systematic search and inclusion of studies.

Of the included studies, 8 included data from African countries (3 from Sierra Leone, 3 from Liberia, 1 from Ghana, and 1 from Senegal), 10 included data from North America (6 from Canada and 4 from USA), 5 were conducted in Australia, 2 were conducted in the UK, and one further study included data from the UK and Australia combined. Most papers (22/29) addressed general issues around social distancing or dealt with multiple explicit measures, among which quarantine was the most dominant one, 3/29 papers exclusively addressed quarantine and 4/29 papers focused on school closures or school-based social distancing while also addressing general concerns. A total of 2199 participants were interviewed or participated in focus group discussions (FGDs), with one study not explicitly reporting the number of participants. Table 1 shows a full list of included studies with information on key characteristics. With regards to study quality, we found that generally, few papers report the reasoning behind data collection and analytical methods used. Only 3 out of 29 reports included indications of reflexivity. In spite of flaws in reporting, all studies provided valuable insights, and appeared to have been conducted appropriately. None of the studies that met the inclusion criteria were excluded based on poor quality. Instead, quality issues were considered when evaluating confidence in review findings using GRADE-CERQual.

**Table 1:** List of included studies. Information on sample, study design, setting, aims and the CASP quality rating (QA Casp) is presented for each study; for the Quality Appraisal, Y = yes, N = no, and U = unclear. The order of criteria follows the order in the tool (1. clear statement of aims, 2. appropriate qualitative methodology, 3. appropriate research design, 4. appropriate recruitment, 5. appropriate data collection, 6. reflexivity, 7. ethical considerations, 8. rigour of data analysis, 9. clarity of statement of findings, 10. value of research). FGDs = Focus group discussions, HCWs = Healthcare workers,

| Study | Setting | Participants | Study Design | Aims | QA (CASP) |
| --- | --- | --- | --- | --- | --- |
| Abramowitz et al. 2015 (19) | 15 communities in Liberia | 386 community leaders | 15 Focus Group Discussions (FGDs) | To identify "mechanisms for community-based response" to a West African Ebola epidemic | Y/Y/U/U/U/<br>N/Y/U/Y/Y |
| Adongo et al. 2016 (20) | 5 regions in Ghana | 235 participants in discussions + 40 community leaders | 25 FGDs and 40 in-depth interviews | To identify "socio-cultural factors that may influence the prevention and containment of EVD in Ghana" | Y/Y/Y/Y/U/<br>N/Y/U/Y/Y |
| Adongo et al. 2016 (21) | 5 regions in Ghana | 235 participants in discussions + 40 community leaders | 25 FGDs and 40 in-depth interviews | To explore "community knowledge and attitudes about Ebola and its transmission" | Y/Y/Y/Y/U/<br>N/Y/U/Y/Y |
| Baum et al. 2009 (22) | Michigan, USA | 37 members of the public | 4 FGDs | "To evaluate public willingness to accept and comply with social distancing measures likely to be imposed during a pandemic" | Y/Y/U/Y/U/<br>N/N/U/Y/Y |
| Braunack-Mayer et al. 2010 (23) | Adelaide, Australia | 1 forum with 9 participants and 1 forum with 12 participants | 2 deliberative forums (each on different questions) | "To elucidate community perspectives on some of the strategies proposed for pandemic planning in Australia" | Y/Y/Y/U/Y/<br>N/Y/U/Y/Y |
| Braunack-Mayer et al. 2013 (24) | One Australian city | 4 principals, 25 staff, 14 parents and 13 students from 5 schools | Interviews | "To examine the implementation of school closures as a strategy to manage a local outbreak of a pandemic strain of influenza" | Y/Y/U/Y/U/<br>N/Y/U/Y/Y |
| Caleo et al. 2018 (25) | One village in Sierra Leone | 20 households and 18 key informants | Semi-structured interviews | "Understanding transmission dynamics and community compliance with control measures over time" | Y/Y/Y/Y/U/<br>N/Y/U/Y/Y |
| Cava et al. 2005 (26) | Toronto, Canada | 21 individuals with experience of quarantine | Semi-structured interviews | "To explore the experience of home quarantine during the SARS outbreak in Toronto in 2003." | Y/Y/Y/Y/U/<br>N/Y/U/Y/Y |
| Cava et al. 2005 (27) | Toronto, Canada | 21 individuals with experience of quarantine | Semi-structured interviews | "To explore the experience of being on SARS quarantine with a focus on the relationship between perceived risk of contracting SARS and reported compliance with the quarantine order and protocols" | Y/Y/Y/Y/U/<br>N/Y/U/Y/Y |
| Davis et al. 2015 (28) | Melbourne, Sydney and Glasgow | 116 purposively chosen participants | 57 interviews and 10 FGDs | "To identify how members of the general public respond to pandemic influenza" | Y/Y/Y/Y/Y/<br>N/Y/Y/Y/Y |
| Davis et al. 2014 (29) | Melbourne, Sydney and Glasgow | 116 purposively chosen participants | 57 interviews and 10 FGDs | "To conceptualise how publics take on the threat of a global respiratory pathogen" | Y/Y/Y/Y/Y/<br>N/U/Y/Y/Y |
| Davis et al. 2011 (30) | Australia | 4 policymakers (and documents) | Interviews (and document study) | "Understanding how pandemic control's assumptions regarding the general public take the specific form they do" | Y/Y/U/U/U/<br>U/Y/Y/Y/Y |
| Desclaux et al. 2017 (31) | Senegal | 43 contact subjects and 27 monitorers | Semi-structured interviews and context data | "Analysing contact cases' perceptions and acceptance of contact monitoring" | Y/Y/U/U/U/<br>U/Y/U/Y/Y |
| DiGiovanni et al. 2004 (32) | Toronto, Canada | 35 participants in unstructured interviews, six FGDs | Unstructured and structured interviews, FGDs | "To cull lessons from Toronto's experiences with large-scale quarantine during the outbreak of Severe Acute Respiratory Syndrome in early 2003" | Y/Y/U/U/U/<br>N/U/U/Y/Y |
| Faherty et al. 2019 (33) | USA | 158 individuals | 36 FGDs | "To present perspectives of school and preparedness officials on the feasibility of implementing a range of social distancing practices" | Y/Y/U/Y/U/<br>N/Y/U/Y/Y |
| Gray et al. 2018 (34) | One urban, one rural location in Sierra Leone | 65 participants (survivors, community members, HCWs) | In-depth interviews (and field notes) | To gain "an understanding of community interactions with the Ebola response" | Y/Y/Y/Y/Y/<br>N/Y/Y/Y/Y |
| Henrich and Holmes 2011 (35) | Vancouver, Canada | 85 people from different groups such as parents, students, HCWs | 11 FGDs | "To begin understanding the communication needs of the public and health care workers" | Y/Y/Y/Y/Y/<br>N/Y/U/Y/Y |
| King et al. 2018 (36) | Sydney, Australia | 42 people in interviews, 431 quantitative surveys | Semi-structured interviews (and quantitative survey) | "To explore what information sources parents trusted and used to obtain information about pH1N1, during both the acute and post- pandemic phase" | Y/Y/Y/Y/Y/<br>N/Y/U/Y/Y |
| Kinsman et al. 2017 (37) | One urban, one rural location in Sierra Leone | 118 people in FGDs and 24 key informants | 16 FGDs and 24 in-depth interviews | "Development of a set of action-able Ebola messages that res-ponded directly to (the com-munity's) needs and concerns" | Y/Y/Y/Y/Y/<br>Y/Y/U/Y/Y |
| Leung et al. 2008 (38) | Toronto, Canada | 19 homeless service providers, clinicians and public health officials | Semi-structured interviews | "To identify the unique challenges related to homeless people that arose during the SARS outbreak and to outline lessons learned that could contribute to planning for future outbreaks" | Y/Y/Y/Y/Y/<br>N/Y/U/Y/Y |
| Mitchell et al. 2014 (39) | Delaware, USA | 48 participants in FGDs + 9 in interviews | FGDs and interviews | "To explore attitudes and behaviours on campus during the first known university outbreak of A(H1N1)pdm09 in the United States, to examine adherence to protective measures, and to determine willingness to follow such recommendations during future influenza outbreaks" | Y/Y/Y/U/Y/<br>N/U/U/Y/Y |
| Morrison and Yardley 2009 (40) | Southern England | 31 participants | 8 FGDs, 1 interview | "To develop a detailed understanding of the interrelated factors which might support or inhibit the adoption of ... infection control measures" | Y/Y/Y/Y/Y/<br>N/Y/Y/Y/Y |
| Pellecchia et al. 2015 (41) | 2 counties in Liberia | 432 participants in FGDs and 30 for interviews | 45 FDGs and 30 semi-structured interviews | "To assess Liberian community perspectives on State-imposed Ebola public health and outbreak containment measures" | Y/Y/Y/Y/U/<br>Y/Y/U/Y/Y |
| Pellecchia 2017 (42) | Montserrado and Grand Cape Mount, Liberia | Unclear | Participant observation, FGDs and in-depth interviews | "To offer an ethnographic account and some reflections on quarantine and the events surrounding its implementation" | Y/Y/Y/U/U/<br>N/N/U/Y/Y |
| Rosella et al. 2013 (43) | 5 Canadian provinces | 40 interviewees (PH officials and scientific advis.), 76 policy docs. | Semi-structured interviews (and document analysis) | "To analyse the public health decision-making process and identify the factors that influenced the uptake and application of evidence for public health policy decisions" | Y/Y/Y/Y/Y/<br>N/Y/Y/U/Y |
| Seale et al. 2012 (44) | Sydney, Australia | 20 university students | Semi-structured interviews | "To measure the perceptions and responses of staff and students at our University (to the 2009 H1N1 pandemic)" | Y/Y/Y/Y/Y/<br>N/Y/U/Y/Y |
| Smith et al. 2012 (45) | Vancouver, Winnipeg and Saint John (Canada) | 17 participants | 3 FGDs | "To present ... findings from the town hall discussions on restrictive measures with the view of further bolstering our empirical understanding of the justifiability of using restrictive measures to achieve public health goals" | Y/Y/Y/Y/Y/<br>N/Y/Y/Y/Y |
| Teasdale and Yardley 2011 (46) | Hampshire, UK | 48 participants | 11 FGDs | "To explore people's beliefs, perceptions, reasoning, and the emotional and contextual factors that may influence responses to government advice for managing flu pandemics" | Y/Y/Y/Y/Y/<br>N/Y/U/Y/Y |
| Uscher-Pines et al. 2007 (47) | Philadelphia, USA | 17 pandemic planners | In-depth interviews | "To collect information related to planning needs and challenges faced by institutions of higher learning, to guide future preparedness activities and the development of specific recommendations for universities" | Y/Y/U/Y/U/<br>U/N/U/U/Y |

### Barriers to the implementation of social distancing measures

Barriers and facilitators identified in the included studies can broadly be categorised into two main types. A full list of concepts with examples of first and second order themes is provided in supplementary table 1.

### Psychological, psychosocial and sociological influences

The first category of barriers comprises individual- and community-level factors. Study participants frequently reported a lack of trust in government and authorities as an important barrier to adherence. (22,24,25,31,34) As a focus group participant in one of the studies described,

> *“With the government, we already know, they’re going to know and they’re not going to let us know until a week or two later…”*. (Baum 2009)

Apart from not trusting authorities, for community members, the fear of being stigmatised by their peers as a result of contracting a disease or being in contact with a suspected case was perceived to be a strong barrier to social distancing (21,26,31,32,39,41), as expressed by the following quote from a study in Senegal:

> “*I haven’t worked because during this whole time, they looked at you a certain way because they all knew that I was among those who were held, so it’s not been easy, you know…” (Desclaux 2017)*

In addition to the fear of stigma, the psychological stress induced by uncertainty and measures like quarantine (23,26,31–33,42) was frequently described as a major barrier.

> *“I thought of that movie (Ben Hur) all the time while I was in quarantine because I remember the part of him going and looking for his sister and his mother, where they had that … sickness, leprosy. And they could not be with the rest of the people*… *and that’s how I felt. I was separate from the world*.*”* (Cava 2005)

Study participants further considered people’s lack of knowledge and misconceptions about the disease (21,25,27,37,46), inconsistencies between personal experience and information received (19,25,27,45), a perceived lack of threat, and the perceived lack of value of interventions (22,27,31,40,44,46) to be barriers to social distancing adherence:

> “I would have to weigh the amount of risk vs. the potential for panic and for there to be a backlash against the kinds of rules that are being instituted. … there’s a balance between over-reacting and under-reacting to a situation …” (Smith 2012)

Many study participants described a perceived lack of community collaboration (22– 24,27,28,31,32,34,40) as an important barrier:

> *“But then I would think if I was to do this, the next, the next person isn’t, why should I blow out the stops”* (Morrison Yardley 2009)

Feelings of solidarity on the other hand were described as crucial to overcome this barrier:

> *“We’re all trying to be good citizens. And we’re all trying to help, you know, other people by making sacrifices like being in quarantine*.*”* (Cava 2005)

Further influences that could become barriers were the inability to work and resulting financial hardship (23,26), dependence on social networks and support systems (22,23,41), social-cultural norms and perceived gender roles (20,25) as well as practical reasons like wanting or having to care for others (40,46).

### Perceived shortcomings in governmental and authority action

With regards to governmental and authority action, study participants lamented the lack of community involvement. (22,24,30,33,41,43,45)

> *“Listen to the average citizens. If there are task forces, citizens should be on each task force*.*”* (Baum 2009)

They further criticised the insufficiency of emotional, financial or material support and cited this as a key reason for non-adherence. (22,23,25,26,31,32,34,38,41,42,45,47)

> *“We had no food at the start. They should have given us food like they did in other households at the end*.*”* (Caleo 2018)

Poor communication was identified as one of the most important factors affecting implementation and adherence to measurements. This included a lack of guidance and ambiguous messaging (19,22,24,26,31,47), as demonstrated by the following quote from a study participant:

> *“I sometimes felt as if I was getting mixed messages. And even the ladies who called from Public Health* … *one I believe said when you’re by yourself you didn’t need the mask. But then the other one said, well no, you have to keep it on all the time*.*”* (Cava 2005)

Further aspects of poor communication cited by study participants were unsuitable messages (19,24,26,35–38), a lack of credibility (24,27,34,39) as well the inadequacy of timing (19) and channels of communication (22,29,32,37,45). Inadequate preparedness (30,32,38), the lack of legislation and penalties (23,25,27,34,45) and authorities’ failure to take equity into account (24,26,38) were additional barriers brought up by participants in a range of settings.

### How to facilitate implementation of social distancing measures

Based on these barriers, and with due consideration of enablers of social distancing described in the included studies, the review identified 25 themes that can be addressed to improve the implementation of social distancing. These themes belong to one of the two broad categories described above. Additionally, because of the richness and coherence of data that support them, themes around communication are listed in a distinct sub-category (see table 2).

**Table 2:** Summary table of review findings and confidence assessment using the GRADE-CERQual approach. C = countries, ET = epidemic threats

| Review Finding | Contributing Studies (N) | Confidence (CERQual) | Notes on confidence rating |
| --- | --- | --- | --- |
| **1. Psychological / psychosocial / sociological factors** |  |  |  |
| **Avoiding stigma:** Efforts should be made to avoid stigma in order to lower the psychosocial cost of adherence. | N = 6<br>(21,26,31,32, 39,41) | moderate | Evidence from 5 countries (C) and 3 different epidemic threats (ET). High relevance and coherence, minor concerns around adequacy, and minor methodological concerns for some of the studies. |
| **Emotional support:** Addressing the psychological burden of quarantine and other SD measures is an important enabler of adherence. | N = 4<br>(23,26,32,33) | moderate | Evidence from 3 C and 2 ET. High relevance and coherence, minor concerns around adequacy, and minor methodological concerns for some of the studies. |
| **Building trust:** A lack of trust in government and authorities impedes people's adherence to SD and should be prevented through constant trust-building efforts. | N = 5<br>(22,24,25,31, 34) | moderate | Evidence from 4 countries and 3 ET. High relevance and coherence, minor concerns around adequacy, and minor methodological concerns for some of the studies. |
| **Solidarity:** Feelings of solidarity, social responsibility and the presence of community collaboration can be important in increasing acceptability of and adherence to measures. | N = 9 (22–24,27,28,31, 32,34,40) | moderate | Evidence from 7 C and 3 ET. High relevance and coherence, minor concerns around adequacy, and minor methodological concerns for some of the studies. |
| **Perceived threat & value of interventions:** The perception of threat and the perception of interventions being effective ways to battle that threat are important for the adherence to measures. | N = 6<br>(22,27,31,40,44,46) | moderate | Evidence from 5 C and 3 ET. High relevance and coherence, minor concerns around adequacy, and minor methodological concerns for some of the studies. |
| **Alignment of messaging and lived experience:** It should be considered that people's personal experience being different from the depiction of the situation by media and authorities is a barrier to SD adherence. | N = 4<br>(19,25,27,45) | low | Evidence from 3 C and 2 ET. High relevance and coherence, some concerns around adequacy, and minor methodological concerns. |
| **Expecting unintended consequences:** With regards to school closures, one problem with regards to social distancing is the compensatory increase in outside-of-school social activities. | N = 2 (39,43) | low | Evidence from 2 C regarding pandemic influenza. High relevance and coherence, some concerns around adequacy, and minor methodological concerns. |
| **Accounting for life circumstances:** It should be considered that practical and circumstantial reasons, like the need to care for others, the need to access services or simply the lack of space can be barriers to adherence to SD. | N = 2 (40,46) | very low | Evidence from 1 C regarding pandemic influenza. High relevance and coherence, major concerns around adequacy, and minor methodological concerns. |
| **Addressing social norms:** Perceived gender roles and habits like handshaking should be addressed directly because they can be barriers to the implementation of Social Distancing (SD) measures. | N = 2 (20,25) | very low | Evidence from 2 C regarding Ebola. High relevance and coherence, major concerns around adequacy, and some methodological concerns. |
| **2.1 Government / authority factors** |  |  |  |
| **Government support:** It is important that governments and authorities provide support for people who adhere to social distancing so that no (few) negative consequences stem from adherence. Different kinds of support should be provided, e.g. emotional, medical, material, financial support. | N = 12<br>(22,23,25,26,31,32,34,38,41,42,45,47) | high | Evidence from 6 C and 3 ET. High coherence, adequacy and relevance. Minor methodological concerns compensated by high-quality studies. |
| **Community involvement:** Involving communities is critical in the planning and response phases of epidemics. | N = 7<br>(22,24,30,33,41,43,45) | moderate | Evidence from 4 C and 2 ET. High relevance and coherence, minor concerns around adequacy, and minor methodological concerns for some of the studies. |
| **Appropriate legislation:** The implementation of legislation and the use of penalties appear to be acceptable and can increase adherence to SD measures. | N = 5<br>(23,25,27,34,45) | moderate | Evidence from 3 C and 3 ET. High relevance and coherence, minor concerns around adequacy, and minor methodological concerns for some of the studies. |
| **Preparation is key:** In order to enable implementation, pandemic plans should be sufficiently detailed and actionable. Preparedness can improve adherence to SD. An example of this are online learning capabilities of schools and universities. | N = 5<br>(24,30,32,38,47) | moderate | Evidence from 3 C and 2 ET. High relevance and coherence, minor concerns around adequacy, and minor methodological concerns for some of the studies. |
| **Continuous communication:** Authorities should provide constant updates and inform the public and especially those affected by social distancing measures like quarantine of new developments | N = 3<br>(22,26,31) | low | Evidence from 3 C and 3 ET. High relevance and coherence, some concerns around adequacy, and minor methodological concerns. |
| **Balancing different interests:** Where possible, social consequences of transmission control should be considered, and breaking social networks and support systems should be avoided. | N = 3<br>(22,23,41) | low | Evidence from 3 C and 2 ET. High relevance and coherence, some concerns around adequacy, and minor methodological concerns. |
| **Taking equity into account:** Governments and authorities should pay attention to equity issues which can be important influences on adherence to SD. | N = 3<br>(24,26,38) | low | Evidence from 2 C and 2 ET. High relevance and coherence, some concerns around adequacy, and minor methodological concerns. |
| **Being clear and transparent:** Clear statements from public health authorities enable the implementation of measures like school closures. | N = 3<br>(33,43,47) | very low | Evidence from 2 C regarding pandemic influenza. High relevance and coherence, major concerns around adequacy, and some methodological concerns. |
| **Providing constant reminders:** The public should be reminded of necessary measures in order to avoid a regression to previous norms | N = 2 (31,40) | very low | Evidence from 2 C and 2 ET. High relevance and coherence, major concerns around adequacy, and some methodological concerns. |

| **2.2 Communication-related factors** |  |  |  |
| --- | --- | --- | --- |
| **Good communication is critical:** Messaging and messengers should be credible, and people call for experts to be on the forefront of communication with the public. There is mistrust against the media. Communication should be transparent, timely, clear and uniform, and it should acknowledge uncertainty and the need for adaptation to changing circumstances. | N = 14<br>(24,26,27,29,30,32–40,45,46) | high | Evidence from 7 C and 3 ET. High coherence, adequacy and relevance. Minor methodological concerns compensated by high-quality studies. |
| **Improving knowledge and addressing beliefs:** Providing knowledge and battling misconceptions about the disease might be valuable ways to increase adherence to SD measures. | N = 4<br>(21,29,44,46) | moderate | Evidence from 4 C and 2 ET. High relevance and coherence, minor concerns around adequacy, and minor methodological concerns for some of the studies. |
| **Relevance and context-specificity of messaging:** Information provided to the public should be context specific and relevant to people's lives. | N = 7<br>(19,24,26,35–38) | moderate | Evidence from 5 C and 3 ET. High relevance and coherence, minor concerns around adequacy, and minor methodological concerns for some of the studies. |
| **Tailoring messages to recipients' needs:** Messaging should be tailored to the diverse communities of recipients. "One size fits all" approaches should be avoided. | N = 4 (35–38) | moderate | Evidence from 3 C and 3 ET. High relevance and coherence, minor concerns around adequacy, and minor methodological concerns for some of the studies. |
| **Doctors contributing as trusted messengers:** Doctors, especially family physicians can act as highly trusted and influential messengers in the response. They should be a) prepared, e.g. by having email addresses of their patients etc., and b) be included during the pandemic as central conveyors of information. | N = 2 (35,36) | very low | Evidence from 2 C focussing on pandemic influenza. High relevance and coherence, major concerns around adequacy, and some methodological concerns |
| **Direct two-way communication:** Direct communication between e.g. schools and public health authorities increases effectiveness of SD implementation. | N = 2 (24,33) | very low | Evidence from 2 C focussing on pandemic influenza. High relevance and coherence, major concerns around adequacy, and methodology |

Data from the studies included in this review indicate that it is important to address stigmatisation and the psychological burden of measures like quarantine. (21,23,26,31–33,39,41) Building trust in government and authorities as well as promoting confidence in the implemented measures are further opportunities for improvement (22,24,25,27,31,34,40,44,46). Addressing solidarity, social responsibility and community collaboration promotes adherence and is a critical element of the response (22–24,27,28,31,32,34,40).

With regards to actions taken by governments and authorities, the most central theme that emerged from the analysis of data in this review is the importance of providing support (emotional, medical, material, and financial) for people who adhere to social distancing, so that no or few negative consequences stem from adherence (22,23,25,26,31,32,34,38,41,45,47). Governments and authorities need to include the community in the planning before and in the response during epidemics (22,24,30,33,41,43,45). Furthermore, the implementation of legislation and the use of penalties appear to be an acceptable means of increasing adherence to social distancing measures (23,25,27,34,45).

Ultimately, the most central theme identified across studies is the critical importance of good communication (24,26,27,29,30,32–40,45,46). Messages and messengers should be credible. Many study participants reported a mistrust of the media and instead asked that scientific experts be at the forefront of communication with the public. With regards to the dynamics of communication, there is broad coherence across the included studies regarding the importance of acknowledging uncertainty and the need for adaptation to changing circumstances. Messages should be tailored to the diverse communities of recipients (35–38), and information should be context specific and relevant to people’s lives. Further important aspects identified were transparency, good timing, clarity and uniformity (19,24,26,35–38).

Table 2 displays a complete list of review findings. Each finding is presented alongside its corresponding GRADE-CERQual confidence rating and the studies that contribute to it.

## Discussion

To the authors’ knowledge, this is the first systematic qualitative review focussing on the implementation of social distancing measures. The review identifies a list of 25 factors that can potentially affect implementation of and adherence to social distancing measures. These factors can broadly be summarised under the themes of individual- or community-level psychosocial factors on the one hand, and government or authority factors on the other. While in reality there are likely many complex relationships between the different factors influencing social distancing acceptability, the schematic depiction in figure 2 (below) may be a useful conceptual way to understand what determines people’s willingness to adhere to social distancing.

**Figure 2:**
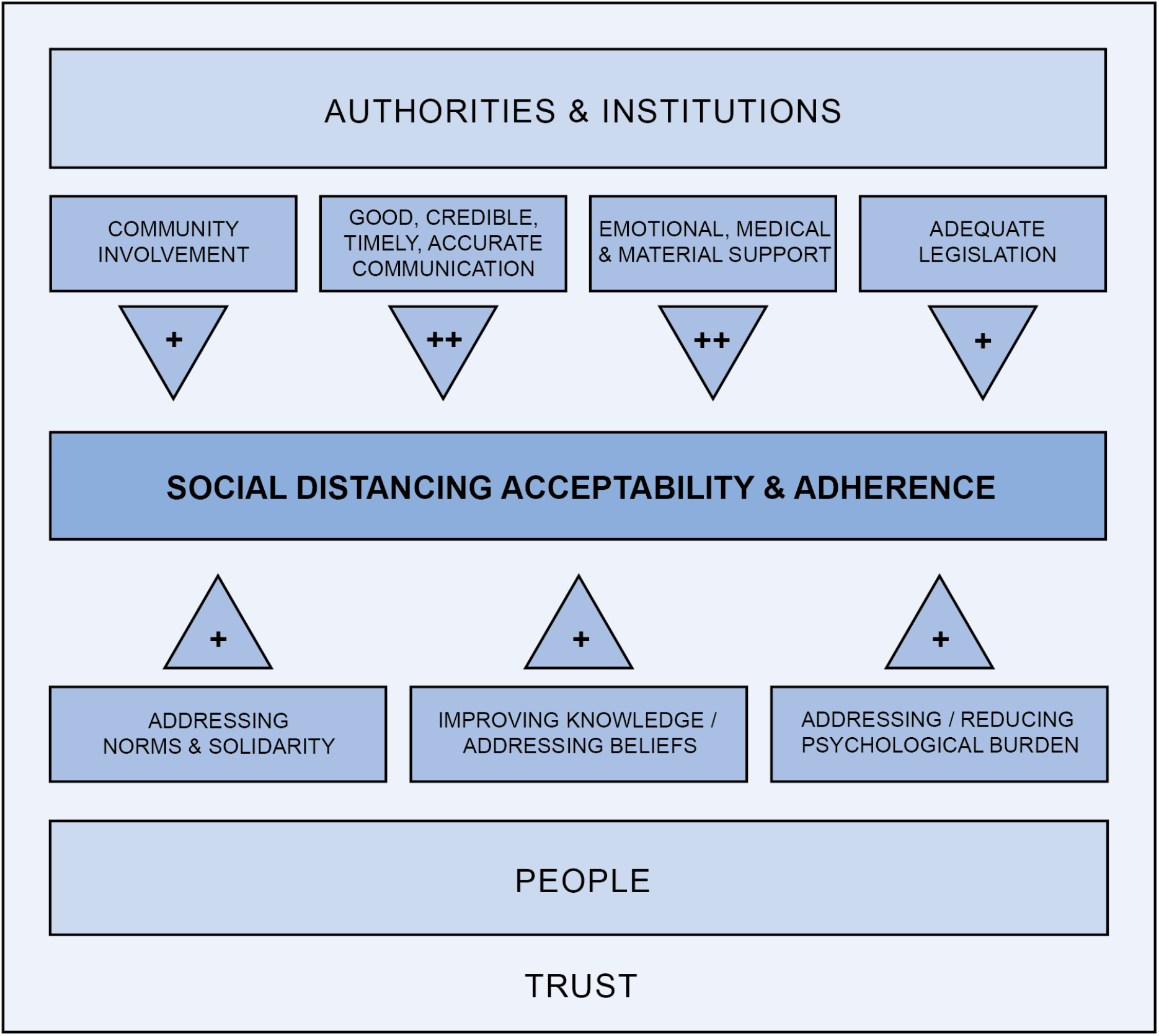
Factors influencing the acceptability of and adherence to social distancing measures. “+” indicates moderate confidence, and “++” indicates high confidence in the factor being an important enabler of social distancing acceptability and adherence.

Where aspects of social distancing were discussed in previous reviews, especially with regards to quarantine and isolation, there is broad agreement on the identified themes, which this review develops further (4,7). Within the studies included in this review, there is broad agreement on the most central barriers and facilitators (as indicated in our summary table 2). Even where there was not enough data to make a high-confidence statement, the review did not find substantial disagreement between the identified studies.

### Implications for policymaking, services, and communication

The review’s findings demonstrate the importance of a comprehensive support system, transparent policies, and sufficient community involvement. They all can contribute to adherence to social distancing measures, and present opportunities for governments to improve the acceptability of mandated measures. The review further indicates that it is critical for policymakers and service providers to recognise the toll measures can take on people. The evidence from the review also shows that preventing stigma, appealing to solidarity, building trust, and making sure that strong support systems are put in place are important in order to alleviate hardship faced by the population that is expected to adhere to social distancing. Finally, effective, transparent, trustworthy communication appears to be a central enabler to the acceptability of and adherence to social distancing measures. Responsible communication should be transparent, timely, clear and uniform, and trusted experts should be at the forefront. Good communication acknowledges uncertainty and the need to adapt to changing circumstances. The evidence also suggests that messaging should be context-specific and relevant to people’s lives. All of these recommendations are concrete and actionable opportunities for policymakers and service providers as well as anyone who communicates with the public.

### Implications for future research

Barriers to and facilitators of social distancing have often been addressed implicitly in the qualitative studies that were identified in this review. Future qualitative research should address implementation more directly.

The systematic searches identified a number of quantitative studies that could complement the review findings in a meaningful way. A mixed methods approach or a future quantitative review may be of value.

Moving forward, findings from this review can inform not only policy implementation but also the research design of future studies to evaluate social distancing measures, their acceptability, feasibility and potential effectiveness. This review further underlines the importance of terminological specificity.

### Limitations of this review

This review has a number of limitations. Firstly, the literature search could have been complemented by searching specific journals and the grey literature.

Most of the studies included in the review have some methodological limitations. The review attempted to account for this in the assessment of confidence for review findings. With regards to whether or not results are broadly representative, included studies were conducted in a limited number of contexts. Geographical areas of the world that are not represented are large parts of Europe and Asia as well as Central and South America. This introduces uncertainty since these measures might be highly settings-dependent.

Importantly, the social distancing scenarios identified in this review are rather short-term. During the coronavirus pandemic, the implementation of social distancing measures has shown to be necessary over a longer period of time which might have a strong influence on adherence. Since no studies had been conducted on the COVID-19 pandemic at the time of the searches, the findings may not be completely representative of the present situation, but they provide an indication of ways to improve the pandemic response. A future review will have to assess new lessons learned and can benefit from the findings established in this work.

Finally, while it is sensible to try and evaluate social distancing broadly, and, as this review has indicated, many findings apply to all aspects of social distancing, it would be worthwhile to pay more attention to the specificities of each social distancing measure, both for evaluating current literature, and for future research.

## Conclusions

This review demonstrates that there is a range of barriers, on different levels, to the implementation of social distancing measures. Some of the key findings are the need for authorities to involve their communities, the need to provide continuous support to those who adhere to social distancing, and the critical importance of good communication. These and many other factors appear to influence acceptability of social distancing and people’s adherence to measures that are necessary for the pandemic response. Policies should be designed with these factors in mind to ensure an effective, ethical and equitable pandemic response. The current situation further calls for high-quality research to better describe mechanisms by which acceptability and implementation of social distancing measures can be improved.

## Data Availability

This is a systematic review. Included studies and data are in the public domain.

