## Supplementary Table 1 for "“Social distancing: barriers to its implementation and how they can be overcome – a rapid systematic review”"

| **First Order (Participant statements)** | **Second Order (synthesis by study authors)** | **Third Order: Barriers to SD** |
| --- | --- | --- |
| **1. Psychological / psychosocial / sociological factors** | | |
| “Until last Monday, I can say that I haven't worked because during this whole time, they looked at you a certain way because they all knew that I was among those who were held, so it's not been easy, you know…” (Desclaux et al. 2017)  “Signs were posted on dorm room doors… : ‘Do not enter if you are sick.’ Sick students felt ostracized.” (Mitchell et al. 2014) | “Because of this broad stigma, three people were unable to resume their jobs when the surveillance ended, with their former employers expressing their fear of the contagion.” (Desclaux et al. 2017)  “A few ill students admitted they did not follow all recommendations, especially staying away from public places and meetings … This was partly related to perceived ostracism.” (Mitchell et al. 2014) | **Stigmatisation** |
| “I thought of that movie (Ben Hur) all the time while I was in quarantine because I remember the part of him going and looking for his sister and his mother, where they had that … sickness, leprosy. And they could not be with the rest of the people... and that’s how I felt. I was separate from the world.” (Cava MA et al. 2005) | “The forum recognised that quarantine and social distancing measures would create social and emotional burdens which would influence people’s willingness and capacity to comply.” (Braunack-Mayer et al. 2010)  “Psychological stress for those in quarantine resulted from social distancing and stigmatization.” (DiGiovanni et al. 2004)  Nearly all the contact persons reported anxiety-induced insomnia during the initial days or ﬁrst week. This anxiety was exacerbated by a lack of information from the follow-up team, who did not mention the very different levels of risk depending on how they had contact with the patient, leaving them to imagine the worst. (Desclaux et al. 2017) | **Psychological and emotional burden (e.g. feelings of anxiety and separateness)** |
| “With the government, we already know, they’re going to know and they’re not going to let us know until a week or two later, after the outbreak has already started so you know, they’re going to get theirs and they’ll be vaccinated. They’re going to make sure their families are taken care of.” (Baum et al. 2009)  “People were hiding symptoms and deaths because they were scared of the camp [EMC]; by the time they were found and the ambulance called, they were already dead.” (Caleo et al. 2018) | “contextual factors encouraged the contact persons’ adherence to monitoring, such as a pre-existing positive appraisal of the health care system and its actors and trust in the national response” (Desclaux et al. 2017)  “They suggested that politicians would do the politically expedient thing rather than what is right in a crisis and that public ofﬁcials may not always convey accurate information to the public.” (Baum et al. 2009)  “Initially, it was difficult for villagers to believe that infection could spread through everyday person-to-person contact. This perception was compounded by a climate of mistrust of authorities…” (Caleo et al. 2018) | **Distrust in the health system and government authorities** |
| “We’re all trying to be good citizens. And we’re all trying to help, you know, other people by making sacrifices like being in quarantine.” (Cava et al. 2005)  "But then I would think if I was to do this, the next, the next person isn't, why should I blow out the stops" (Morrison and Yardley 2009) | “an individualistic approach to pandemic risk may obscure factors that the individual cannot control and, as indicated by the judgement of those who acquired infection, health individualism may be moralising… focus of the healthy on their own health risks (at the expense of others).” (Davis et al. 2015)  “The most important reason for complying was to reduce the risk of transmission to others …, and the majority of interviewees and focus group participants cast this motivation as ‘civic duty’.” (DiGiovanni et al. 2004) | **Lack of solidarity, community collaboration and feeling of social responsibility** |
| “…that right now it’s in some third world country and it may come here. I don’t think that’s going to be good enough. I think there’s going to have to be some indication that it is actually in your own community before you take steps as drastic as shutting down anything.” (Baum et al. 2009)  "I'd say um in terms of a pandemic it makes it sound like it's unstoppable anyway so if you're gonna get it you're gonna get it despite whatever you can do to try and help it it won't stop the pandemic and that's why it's called a pandemic" (Morrison and Yardley 2009) | All groups discussed the need to have compelling reasons to endure the inconvenience or hardships such measures would bring. In light of the expected economic burdens associated with social distancing measures, participants ex-pressed a need to know that threat of a disease was imminent and severe before agreeing to comply with policies that would likely be onerous and disruptive. (Baum et al. 2009)  “To some extent, the degree of reported compliance was related to risk perception and in particular to the person’s assessment of risk for themselves and others.” (Cava et al. 2005) | **Perceived lack of a threat and perceived lack of value of the interventions** |
| “Sensitisation from different sources … started to make sense; symptoms in our loved ones were exactly the same as they were telling us.” (Caleo et al. 2018)  “I would have to weigh the amount of risk vs. the potential for panic and for there to be a backlash against the kinds of rules that are being instituted. … there’s a balance between over reacting and under reacting to a situation ...” (Smith et al. 2012) | “Perceptions of EVD held by the villagers changed when information received from contact tracers and the MSF health promotion team was consistent with what villagers observed in their lives at the community level.” (Caleo et al. 2018)  “Participants stressed that, in order to create an environment for compliance and to justify the use of restrictive measures, measures must be proportional to the risk that is perceived by the public. Furthermore, participants expressed that the actual risk that exists (according to experts) must be balanced with the potential impact of using restrictive measures” (Smith et al. 2012) | **Personal experience being inconsistent with information** |
| “Within my [dormitory] hall, if anything, [students] decided to party.” (Mitchell et al. 2014)  “Yeah. Yeah. We were all, you know, sort of either emailing or talking to each other going, well what do you think? No one’s sick. Right, okay. Reckon it’s safe? We still tried to minimise. It’s not like we’d go to massive sporting events…” (Braunack-Mayer et al. 2013) | “Some students reportedly continued and perhaps intensified their social calendar during the week that activities were cancelled.” (Mitchell et al. 2014)  “Children would congregate in other places, similarly increasing risk of transmission” (Rosella et al. 2013)  “(…) a group determined that they were safe to re-enter the community before the quarantine period was completed. This was based, in part, on rumours circulating through these informal staff cliques that other staff members (including the principal) were already out in the community.” (Braunack-Mayer et al. 2013) | **Compensatory increase in outside-of-school social activities and non-adherent peers** |
| "I think it will still be quite practically hard if like the person that was infected chose just to walk around the house or flat like practically it's really hard to keep three feet away from them, maybe wait till they've walked out the corridor and just things like that" (Morrison and Yardley 2009)  “If I didn’t have enough supplies at home of whatever, then I would have to go out. It wouldn’t make any difference if I had swine ﬂu or not I would just hope that I wouldn’t see anybody, or talk to anybody, …” (Teasdale and Yardley 2011) | “The need or desire to care for ill persons was seen as a major barrier to the implementation of social distancing measures. Indeed, some considered that it would be selfish to "flee" from an infected member of their family purely for self-protection. … A lack of adequate space to maintain social distance was cited as a further practical barrier.” (Morrison and Yardley 2009)  “Common perceived barriers to following the stay home advice were feeling guilty and anxious about missing work and not wanting to let people down, … as well as more practical issues such as needing to go out for essentials (e.g., food, medicines).” (Teasdale and Yardley 2011) | **Practical reasons like wanting or having to care for others, needing to access facilities, lack of space, etc.** |
| “Quarantine cannot work here, it drowned people in fear and economic hardship.” (Pellecchia 2017) | “Among the contact persons, 55 had to interrupt their professional activities with no advanced planning. The social effects of this suspension were primarily economic and secondarily related to roles and social status.” (Desclaux et al. 2017) | **Inability to work and insufficient support** |
| “I don’t know, I think people should have the choice if they need to go to church for whatever reason at this time, this kind of thing and they’re going to make the choice I guess to go out. That might be a place that they need to go.” (Baum et al. 2009) | “State-enforced quarantine broke social networks of solidarity and was implemented with lots of gaps that raised social concern.” (Pellecchia et al. 2015)  Some participants in all four groups shared opposition to mandatory closure of religious organizations during a pandemic. They cited the importance of religious communities for support, for opportunities to worship and pray together during crises. (Baum et al. 2009) | **Dependence on social networks and support systems** |
| “Shaking of hands is very important in our society. If you refuse to accept handshakes you are seen as proud in this community. In funerals, you go round and shake everybody’s hand” (Adongo et al. 2016)  “It’s always a little difficult to tell when you’re moving from, sort of, a cold through the man flu to proper influenza.” (Davis et al. 2015)  “We women do all the washing ofclothes and all the items often used by a sick person, therefore ifthe person should have the condition, it means we will get it” (Adongo et al. 2016) | “Handshaking emerged as one of the everyday communal social norms that may influence Ebola prevention and containment. … To respondents, this was mandatory in some social settings and had deep roots in the culture and behavioral etiquettes of the people …” (Adongo et al. 2016)  “Resistance to specific practices that were perceived as offensive to socio-cultural norms was reported; this resistance continued until the value of such practices was understood.” (Caleo et al. 2018)  “Another important provision on health individualism was the gendered meanings of one’s response to infection. Importantly, the pejorative term ‘man flu’ was used to denote the over-inflation of mild symptoms ... with connotations of questionable masculinity … The uniform implementation of social distancing and other protective measures may therefore be compromised.” (Davis et al. 2015)  “Women showed an intense conviction that they should care for their families, and showed a desire to do so, despite risks to their own health. Childcare, eldercare, home-based healthcare and the graduated triage approach were gen-  dered activities; and home-based care constituted a zone ofrisk for both predominantly female caregivers and their dependents.” (Abramowitz et al. 2015) | **Social norms (like handshaking) and gender roles (like caring responsibilities)** |
| “It's slightly strange to enter a home without sitting down or shaking hands. The ﬁrst few days we tried to follow this rule, but as time passed, we dropped it.” (Desclaux et al. 2017) | “When the fear of transmission receded after several days, the people adhered less to the rationale for social distancing…” (Desclaux et al. 2017)  “The need for memory joggers was advocated with many participants stating that even if they did wish to implement the measures they would most likely forget. This included … posters or campaigns to remind people of the types of behaviours they should be undertaking.” (Morrison and Yardley 2009) | **Regression to the norm / Lack of constant reminders** |
| “We thought it was a curse; some people thought that it was some kind of traditional medicine that was being thrown on them.” (Caleo et al. 2018) | “People’s own personal beliefs about ﬂu transmission and vaccinations played an important role in appraising and responding to the government recommendations. A common belief to emerge from the focus groups was that ﬂu transmission is principally airborne, which prompted doubts about the effectiveness of the recommendation to stay home if symptomatic.” (Teasdale and Yardley 2011)  “Reduced misgivings and doubt about Ebola were crucial to influencing attitudes toward control measures. This change likely occurred once the health messages given to the community mirrored their reality.” (Caleo et al. 2018) | **Lack of knowledge and presence of misconceptions about disease** |
| **2.1 Government- or authority-level barriers** | | |
| “We had no food at the start. They should have given us food like they did in other households at the end.” (Caleo et al. 2018)  “And I thought, what do they expect? Now I always keep a supply of food. But I thought what if someone is on their own, and they can’t get food, they can’t get out. They’re on their own. Where is this help?” (Cava MA et al. 2005) | “The quality of human interactions with those implementing control measures was important, in that expressions of dignity, respect, and compassion were key components for positive engagement.” (Gray et al. 2018)  “Study participants, both those under quarantine and neighbours, explained how food, water and other items were only intermittently distributed by the implementing agencies, creating harm and forcing residents to disobey the imposed isolation.” (Pellecchia et al. 2015) | **Insufficient supporting systems (emotional, financial, material)** |
| “Focus groups are important. Listen to the average citizens. If there are task forces, citizens should be on each task force.” … “Listen to the people. Groups like this are important. The public have [sic] well meaning opinions that the policy makers may not know. Listen to the people.” (Baum et al. 2009) | “Local leaders who participated in the research reported a lack of involvement in the decisional forums and felt the form of the State-enforced quarantine did not comply with local communities’ dynamics. In addition, spiritual leaders were not consulted, despite their strong influence in the communities.” (Pellecchia et al. 2015)  “(People stressed the need for) opportunities for public input into pandemic response planning.” (Baum et al. 2009) | **Lack of community involvement in the planning and response** |
| “But we had to follow the law we had to pay 500,000 Leones if there was a sick person found in the house.” (Caleo et al. 2018)  “Well ‘cause of the fines and stuff. ... [I]f they find out you were gone it’s like $5,000 from you. Like you know what? We won’t take the risk.” (Cava et al. 2005) | “Implementation of the by-laws on travel and penalties for not reporting cases supported the understanding of the severity of the outbreak by villagers, and helped them accept that control measures were intended to protect and help the community.” (Caleo et al. 2018)  “Nearly all participants (11/12) agreed that warnings, cautions and fines for infringements were appropriate; most (9/ 12) considered that monitoring the activities of those who had infringed quarantine or social distancing requirements was acceptable” (Braunack-Mayer et al. 2010) | **Lack of Legislation and penalties** |
| “We were the first school, luckily for us we are already an on-line school. I worked at another school for 12 months while I was here, I don’t know how they would have done it because there was no student-teacher email” (Braunack-Mayer et al. 2013)  “Why aren’t kids being taught about public health in schools so that in fact it normalises in many ways that these alerts will come from time to time and [that] we’ll have to change the way we act as social beings to protect ourselves and to protect our communities.” (Davis et al. 2011)  “Well we have a generic plan for those sort of things......, and we swung that plan into action.....But that’s more of a disaster plan and this really in a sense wasn’t disaster but it was an issue that had to be managed and contained.. . on an ongoing rolling basis.. .” (Braunack-Mayer et al. 2013) | “Some schools used email and websites to ensure students particularly final year students, continued with schoolwork and had access to teachers during the quarantine period.” (Braunack-Mayer et al. 2013)  “In the event that courses would be cancelled, many schools had plans to improve their distance education capabilities and train faculty on such systems. Increasing server capacity and digitizing classroom materials in advance were strategies aimed to facilitate online learning.” (Uscher-Pines et al. 2007)  “However, the potential extended duration of a pandemic will likely require planning beyond that traditionally done for all hazards” (Uscher-Pines et al. 2007)  “Initial procedures instituted by schools at the outset of the closure were often based on generic ‘school emergency’ plans and, given the lack of prior experience, principals suggested that they had to “make things up” with guidance from government officials.” (Braunack-Mayer et al. 2013) | **Lack of or inadequate, unspecific preparedness (e.g. online preparedness for schools)** |
| “The official line was we refer to the government website ... and a lot of the information on there was quite ambiguous... And it was really putting the onus back onto the individual to make a judgement, as was the case with me.” (Braunack-Mayer et al. 2013)  “As some respondents noted, one message said, “Don’t touch,” while another said, “Touch, but use plastic gloves.” (Abramowitz et al. 2015)  “I sometimes felt as if I was getting mixed messages. And even the ladies who called from Public Health . . . one I believe said when you’re by yourself you didn’t need the mask. But then the other one said, well no, you have to keep it on all the time.” (Cava et al. 2005) | “Participants shared a desire for accessible and accurate information about limiting contagion. Some complained that they had never received any information about pandemic preparedness or knew of any recent educational efforts within their communities.” (Baum et al. 2009)  “Individuals in quarantine expected Public Health to provide information and reassurance to allay fears and uncertainty.” (Cava MA et al. 2005)  “Although schools expressed a firm commitment to planning for pandemic influenza, we consistently found that they were uncertain as to the appropriate level of detail to incorporate.” (Uscher-Pines et al. 2007) | **Lack of guidance and clear, unambiguous information on adequate behaviour and progress of pandemic** |
| “I asked myself why did it take them 5 days to call me, but then that’s when the light went on and I got scared to death” (Cava MA et al. 2005) | “Continuity of messages and continuity of message delivery was highlighted as a critical issue. Community leaders emphasized the need to engage with communities on a daily basis, and for the messages they were relating to be correct and similar.” (Abramowitz et al. 2015)  “Many expressed frustration at not receiving the quarantine order until several days after exposure.” (Cava MA et al. 2005) | **Inadequate frequency and timing of updates** |
| - | “Uptake and acceptance for most participants was motivated by … the credibility and availability of resources” (Gray et al. 2018)  “The government’s failure to adequately convey a clear rationale for the measures undertaken tended to reduce the credibility of information and led to widespread misinterpretation of the advice about home isolation.” (Braunack-Mayer et al. 2013)  “Many participants expressed doubts about the credibility of information they received from the public health department. These doubts affected their perception of risk.” (Cava et al. 2005) | **Lack of credibility** |
| “we trusted fully with the Department of Health. They inspired a lot of confidence. Again, I think they were fantastic with this, and I was able to ring them and say, look, I’m struggling with this” (Braunack-Mayer et al. 2013) | “The amount of interaction with public health officials varied across the stages of the pandemic but was highest in schools with whole school closures… With a few exceptions, the interactions were positive.” (Braunack-Mayer et al. 2013)  “Strong partnerships across the education and public health sectors are needed to ensure behaviors continue beyond school settings.” (Faherty et al. 2019) | **Insufficient collaboration with and among implementing institutions** |
| “Because we’d spent a lot of years planning and we spent a lot of years engaging with key responders, agencies and the general practice, so whilst we thought we were conﬁdent we had good plans in place, good strategies in place to combat whatever was given to us at the time, it was really a matter of following what we’d developed and agreed to.” (Davis et al. 2011) | “Although schools expressed a firm commitment to planning for pandemic influenza, we consistently found that they were uncertain as to the appropriate level of detail to incorporate. We believe that checklist-type guidance may contribute to this problem, as planning aspects are categorized not by level of detail but by state of completion.” (Uscher-Pines et al. 2007) | **Having a plan but not carrying it out or plans not being actionable** |
| - | “Equity: For example, access to the internet was assumed, but limited, for some students and parents, and public health officials also were not prepared for the provision of information in languages other than English, or to suit varying degrees of health and general literacy. Some schools filled this gap with their own processes for managing language difficulties.” (Braunack-Mayer et al. 2013)  “During the SARS outbreak, members of the general public who had a fever or cough and no known exposure to SARS were told to remain at home until their symptoms had resolved. Health-care providers sometimes failed to recognize the need to adapt this practice when caring for homeless patients…” (Leung et al. 2008) | **Failure to take equity into account** |
| **2.2 Communication-related barriers** | | |
| “The man [index case] came with a letter that said he should be isolated for 21 days. But we didn't understand what ‘isolation’ meant.” (Caleo et al. 2018)  “People will understand the Krio, instead of big English” (Kinsman J. et al. 2017)  “We have heard the messages, but most people do not know how to practicalize them.” (Abramowitz et al. 2015) | “Lack of understanding of complex health messages such as the importance of isolation of those infected.” (Caleo et al. 2018)  “low literacy rate (36% for women and 54% for men [27]) means that text-based posters and billboards will simply be missed by a majority of the population.” (Kinsman J. et al. 2017) | **Overly complex, inaccessible information** |
| “My doctor …He’s an anthroposophical doctor, but I also listen to an alternative radio show on co-op. I think it’s on Thursdays and it’s an alternative radio show that you can call in and there is a homeopathic doctor and … you can just … throw your questions out.” (Henrich and Holmes 2011) | “No single messenger will reach all members of the community: complementary messengers are needed for each setting.” (Kinsman J. et al. 2017)  “Central to combating a pandemic will be to persuade ‘alternative subpopulations’.’ … Because the alternative health care providers are often perceived as adversaries rather than allies, communications with these providers will need to be done in an inclusive and culturally appropriate way.” (Henrich and Holmes 2011) | **Messages not tailored to target group** |
| “take precautions, cover your face… then there wasn’t any-thing else after that… all this scare information to start with, but we haven’t really got anything else that we can run with.” (Cava MA et al. 2005)  “Sensitisation from different sources [MSF/MoHS/ radio] started to make sense; symptoms in our loved ones were exactly the same as they were telling us.” (Caleo et al. 2018) | “…to place themselves on home quarantine and isolate themselves for 10 days. These instructions were obviously impossible for a homeless person...” (Leung et al. 2008)  “Health sensitization efforts continued to emphasize the ‘low-hanging fruit’ of public health communications throughout the response, repeating messages like: ‘What is Ebola? How is it spread? What are the symptoms? … (but) failed to provide the kinds of ‘higher-order’ practical information and training that communities were desperate for, like ‘How do I manage a family of children … in quarantine?’” (Abramowitz et al. 2015)  “Caregiving in all aspects demanded physical contact, but the public health messages regard-  ing physical contact failed to recognize this reality.” (Abramowitz et al. 2015) | **Information not meeting and reflecting people’s realities and needs** |
| “The best is when they hold it [the poster or leaflet] in their hands and move door to door, explaining to the people. But just looking at the picture, it will not be properly understood” (Kinsman J. et al. 2017) | “The messages that can emerge from a two-way process are more likely to be practical, relevant, and actionable, and less likely to focus on the potentially less effective ‘low-hanging fruit’ of health communications, such as those based on slogans like ‘Ebola is real’ or ‘Stop Ebola’. In addition, although two-way communication of this nature requires more capacity and organisation than top-down mass media approaches, it allows for direct community sensitisation, which has the capacity to increase people’s ability to adhere to prevention practices, while also offering the possibility to develop and maintain trust in the messengers.” (Kinsman J. et al. 2017) | **Unidirectional communication** |
| “If you trust that person who carries the message, any message he or she is giving you, you can rely on it and believe in it” (Kinsman J. et al. 2017)  “I want to suggest, in passing on this message, let the religious leaders pass it on to their congregational members, let the youth leaders target the youths, the old target the old, teachers and lecturers pass on it on to their students. With this, it will be nice and the message will pass on to everyone” (Kinsman J. et al. 2017) | “People recognize that mainstream media is a necessary, at least initially, primary source of information. But they do not trust the mainstream media. The solution is for the mainstream media to act as a conduit for information directly from the mouths of lead health officials and experts. Because people trust health information from Web sites of the WHO and CDC, the mainstream media should also publicize links to these sites as well as links to other official regional or national health agency Web sites. Neither the mainstream media itself nor politicians are trusted in a health crisis. The lead health professionals must communicate with the public as directly and as often as possible. They should also speak with one voice.” (Henrich and Holmes 2011)  “However, as with all categories ofputative messengers, the views about their trustworthiness were not universal: some people trusted the medical workers while others did not; equally, some trusted the Chiefs, while others did not. This has important but complex implications when considering who should act as the messenger for a given message.” (Kinsman J. et al. 2017) | **Wrong channels / messengers** |

Supplementary Table 1: First, second and third order themes describing barriers to social distancing. A representative selection of participant quotes (first order themes) and analysis statements from papers (second order themes) are shown on which the identified barriers to social distancing (third order themes) are based.
